# Are vaccines safe in patients with Long COVID? A prospective observational study

**DOI:** 10.1101/2021.03.11.21253225

**Authors:** DT Arnold, A Milne, E Samms, L Stadon, NA Maskell, FW Hamilton

## Abstract

**Introduction:** Although the efficacy of SARS-CoV-2 vaccination to prevent symptomatic COVID-19 is well established, there are no published studies on the impact on symptoms in patients with Long Covid. Anecdotal reports have suggested both a potential benefit and worsening of symptoms post vaccination with the uncertainty leading to some vaccine hesitancy amongst affected individuals.

**Methods:** Patients initially hospitalised with COVID-19 were prospectively recruited to an observational study with clinical follow-up at 3 months (June-July 2020) and 8 months (Dec 2020-Jan 2021) post-admission. Participants who received the Pfizer-BioNTech (BNT162b2) or Oxford-AstraZeneca (ChAdOx1 nCoV-19) vaccine between January to February 2021 were identified and matched 2:1 (in terms of 8-month symptoms) with participants from the same cohort who were unvaccinated. All were re-assessed at 1 month post vaccination (or matched timepoint for unvaccinated cohort). Validated quality of life (SF-36), mental wellbeing (WEMWBS) and ongoing symptoms were assessed at all timepoints. Formal statistical analysis compared the effect of vaccination on recent quality of life using baseline symptoms, age, and gender in linear regression.

**Results:** Forty-four vaccinated participants were assessed at a median of 32 days (IQR 20-41) post vaccination with 22 matched unvaccinated participants. Most were highly symptomatic of Long Covid at 8 months (82% in both groups had at least 1 persistent symptom), with fatigue (61%), breathlessness (50%) and insomnia (38%) predominating. There was no significant worsening in quality-of-life or mental wellbeing metrics pre versus post vaccination. Nearly two-thirds (n=27) reported transient (<72hr duration) systemic effects (including fever, myalgia and headache).

When compared to matched unvaccinated participants from the same cohort, those who had receive a vaccine had a small overall improvement in Long Covid symptoms, with a decrease in worsening symptoms (5.6% vaccinated vs 14.2% unvaccinated) and increase in symptom resolution (23.2% vaccinated vs 15.4% unvaccinated) (p=0.035). No difference in response was identified between Pfizer-BioNTech or Oxford-AstraZeneca vaccines.

**Conclusions:** Receipt of vaccination with either an mRNA or adenoviral vector vaccine was not associated with a worsening of Long Covid symptoms, quality of life, or mental wellbeing. Individuals with prolonged COVID-19 symptoms should receive vaccinations as suggested by national guidance.

## Introduction

It is increasingly recognised that a proportion of patients develop prolonged symptoms following acute SARS-CoV-2 infection, now commonly referred to as Long Covid(^1, 2^). These symptoms are varied but fatigue, breathlessness, myalgia and insomnia are most frequently occurring.

As the immunological basis of Long Covid is unknown, there remains some uncertainty around whether vaccination against SARS-CoV-2 might worsen the associated symptoms^(3)^. Many individuals with Long Covid have expressed hesitancy around receipt of a COVID-19 vaccine for fear of making symptoms worse (Personal Communication with Long Covid Support group); while anecdotal reports of dramatic symptom improvement have also been reported. To our knowledge, no study has attempted to assess this in a cohort of patients with confirmed COVID-19 and a high burden of long-term symptoms.

In this study, we assessed the change in quality of life and symptoms after vaccination amongst a well characterised prospective cohort of patients originally hospitalised with COVID-19 with a significant proportion of persistent symptoms.

## Methods

Between April and May 2020, consecutive patients admitted to a single UK hospital with COVID-19 were prospectively recruited to an observational study and followed up to discharge and at 3 months (June to July 2020) and 8 months (December 2020 to January 2021), as described previously^(4)^. At each clinic, validated quality of life questionnaires (Short Form-36 (SF-36), Warwick and Edinburgh Mental Wellbeing scores (WEMWBS)) were performed, alongside a systematic review of ongoing symptoms.

From this cohort, we identified all participants who had received at least one dose of a COVID-19 vaccine via the National Immunisation Management Service (NIMS). Participants were telephoned approximately 1 month post vaccination (January to February 2021) with quality of life questionnaires and review of symptoms repeated, with specific questions on whether symptoms had improved, stayed the same, or worsened. Participants were only asked to confirm vaccination status after assessment, minimising bias due to perceived association between the assessment and vaccination. Participants were subsequently asked about adverse effects temporally related to the vaccine.

Unvaccinated participants from the same cohort study, matched to the vaccinated participants based on symptomatology and quality of life at 8 months, were identified as a control group and telephoned with the same assessments as above, at the same calendar time post discharge. For brevity the this is referred to as the matched post vaccination time in the unvaccinated group, and post vaccination in the vaccinated group.

T-tests were used to compare 3 and 8-month quality of life and mental wellbeing metrics with the post vaccination metrics, as well as comparing unvaccinated versus vaccinated participants. Median number of symptoms at each time point were compared. Linear models were fitted to formally test for any effect of vaccination on quality of life controlling for 8-month quality of life, age, and gender.

## Results

Forty-four vaccinated participants were assessed at a median of 32 days (IQR 20-41) post vaccination with 22 unvaccinated matched participants. Both groups were highly symptomatic of Long Covid prior to vaccination, with 82% in both vaccinated and unvaccinated groups having at least one persistent symptom at 8-month follow up. Fatigue (61%), breathlessness (50%) and insomnia (38%) were the predominant symptoms. Table 1 shows the baseline demographics.

**Table 1:** Demographics

| Characteristic | Unvaccinated (N = 22) | Vaccinated (N = 44) |
| --- | --- | --- |
| Age | 55 (47, 60) | 64 (54, 73) |
| Male | 13 (59%) | 28 (64%) |
| Female | 9 (41%) | 16 (36%) |
| Diabetes Type 1 | 2 (9.1%) | 0 (0%) |
| Diabetes Type 2 | 3 (14%) | 4 (9.1%) |
| Heart disease | 2 (9.1%) | 11 (25%) |
| Chronic Lung disease | 2 (9.1%) | 14 (32%) |
| SARS-CoV-2 PCR positive during hospitalisation | 18 (82%) | 37 (84%) |
| SARS-CoV-2 Antibody positivity at 3-months | 17 (77%) | 37 (88%) |
| Ongoing Long Covid symptoms at 8-month follow-up | 18 (82%) | 36 (82%) |
| Median (IQR); n (%) |  |  |

Given the vaccine programme in the United Kingdom prioritises older age groups and those with comorbidities, patients who were in vaccinated group were older and more comorbid. The majority were PCR positive during their hospital admission (55/66) with 11 enrolled due to strong clinicoradiological suspicion. 88% of those vaccinated had a positive SARS-CoV-2 antibody test at the 3-month follow-up, compared to 77% in those who are unvaccinated.

### Symptomatology

Symptoms are visualised in Figure 1, stratified between vaccinated and unvaccinated patients, and by difference between 8-months and post vaccination/matched time. The most frequently occurring 8-month symptoms were well matched between the groups allowing for change over time to be examined.

**Figure 1:**
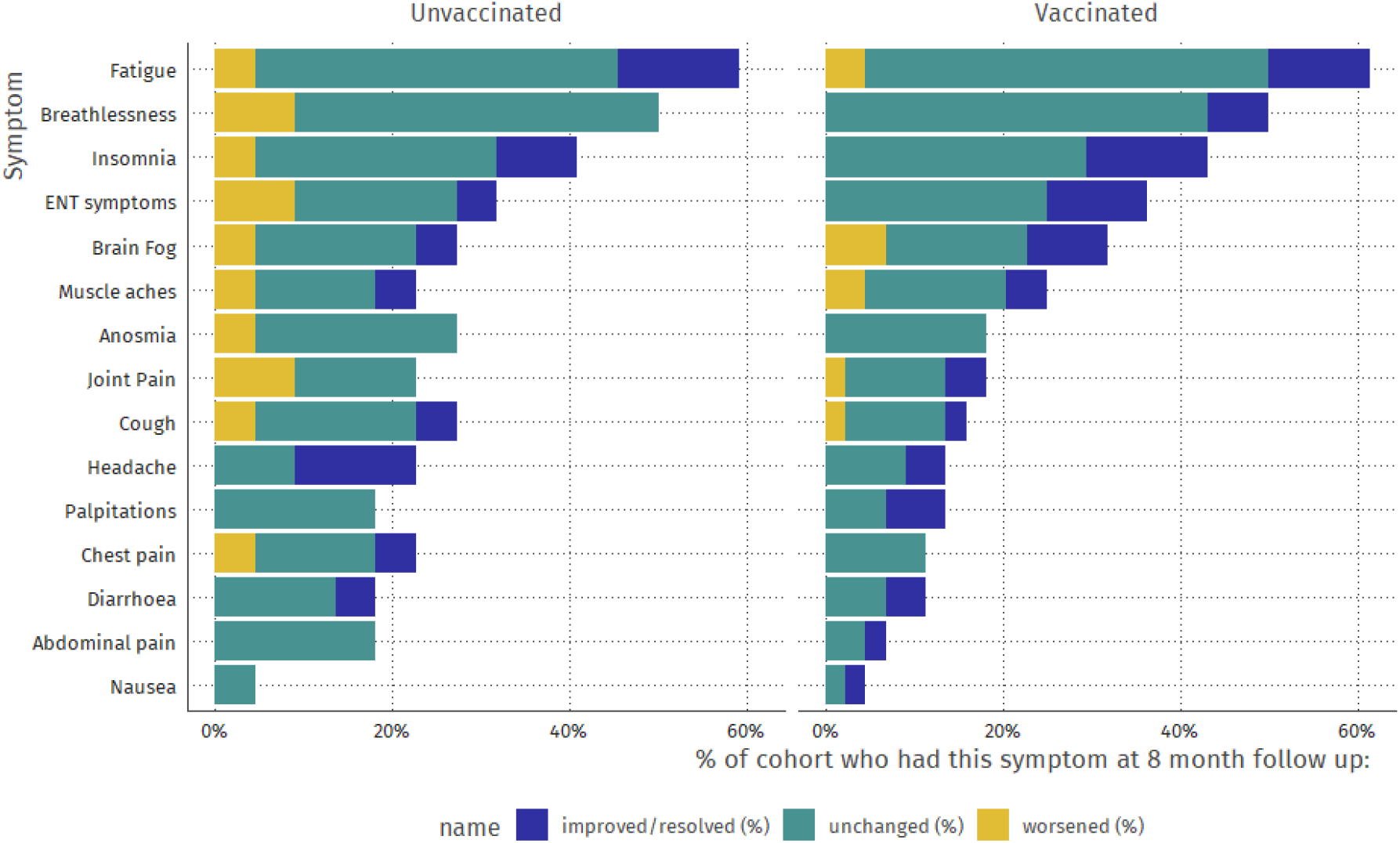
Symptoms at 8-month follow up with change following vaccination (or matched timepoint in unvaccinated group)

There was significant variability in symptoms between timepoints, with patients in both groups having worsening and improvement of symptoms, although most commonly symptoms remained unchanged. A total of 250 symptoms were reported at 8 months across the 66 patients, 91 in the unvaccinated cohort, and 159 in the vaccinated cohort, a median of 4.1 and 3.6 symptoms/patient respectively.

13/91 (14.3%) of symptoms worsened in unvaccinated individuals, compared to 9/159 (5.6%) of vaccinated patients. 64/91 (70.3%) and 113/59 (71.1%) symptoms were unchanged in unvaccinated and vaccinated individuals respectively, with improvements in 14/91 (15.4%) and 37/159 (23.2%) of symptoms in unvaccinated and vaccinated respectively. These differences were statistically significant (p = 0.035, Chi Square).

### Mental and physical composite scores

Figure 2 shows the physical and mental composite scores (PCS and MCS) of the SF-36 score at each time point, with comparisons within each cohort. Unvaccinated and vaccinated participants had similar scores prior to vaccination (p> 0.1 for all comparisons).

**Figure 2:**
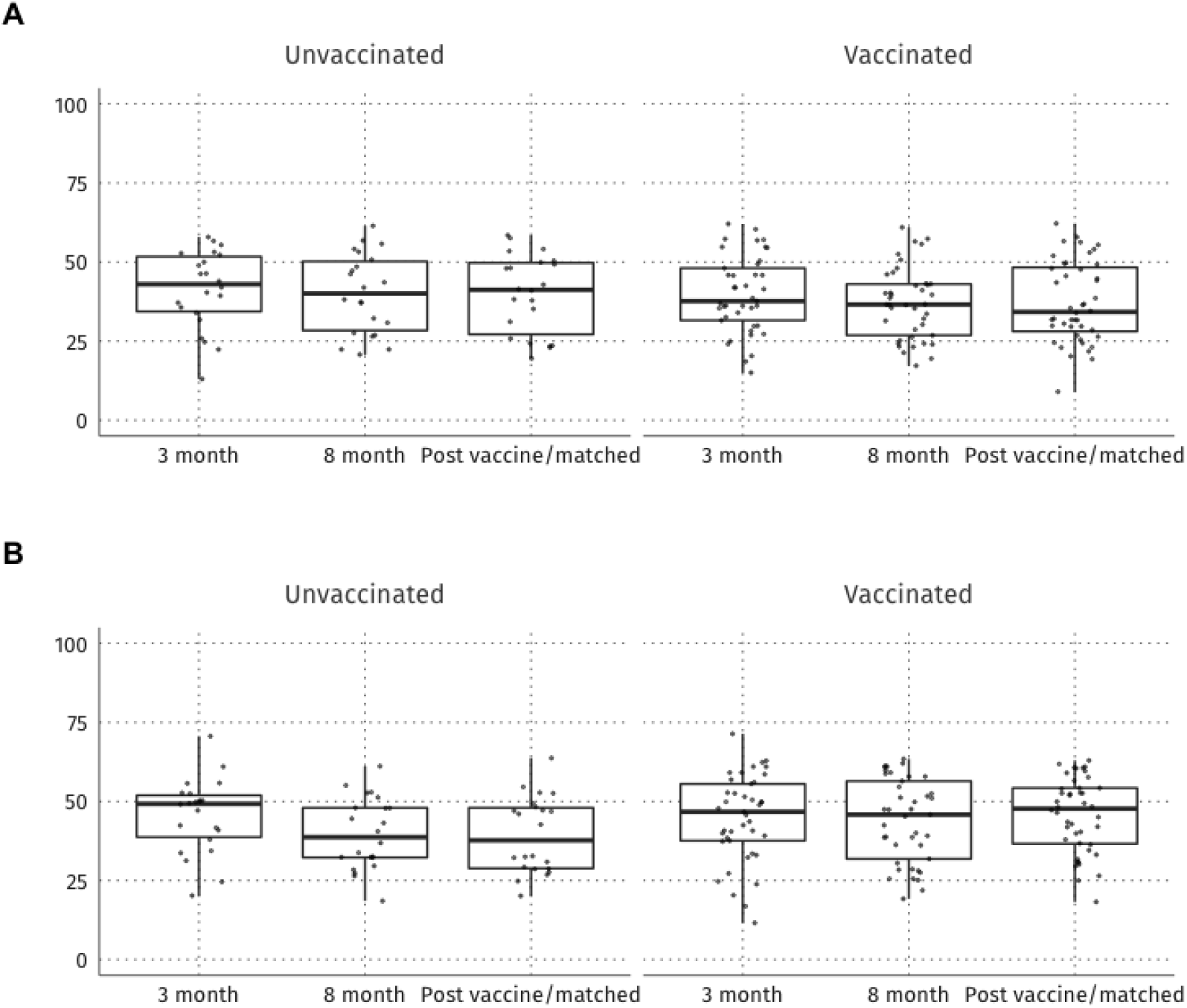
Changes in PCS (a) and MCS (b) over time in vaccinated and unvaccinated cohort.

Post vaccination/matched time point, there was no difference in PCS (median 41; IQR 27-50 in vaccinated, median 34; IQR 28-48 unvaccinated, p = 0.3), however vaccinated participants had higher MCS (median 48; IQR 37-54 vs 38; IQR 29-48, p = 0.039 in vaccinated vs unvaccinated respectively)

This association (higher MCS post vaccination compared to unvaccinated) was not significant in linear models (reported in supplemental tables S1 and S2) once adjusting for 8-month quality of life, age and gender (p = 0.6). In summary, no association (positive or negative) was identified with vaccination status and quality of life.

### Mental wellbeing

Warwick and Edinburgh Mental Wellbeing scores are visualised in Figure 3. Vaccinated participants had broadly similar WEMWBS over the three time points (3 month, 8 month, post-vaccination) with medians (IQR) of 51 (40 – 59), 49 (42 – 57) and 52 (41-61) respectively. In contrast unvaccinated participants had more variability, with scores of 48 (38-54), 45 (36-50), and 54 (46-58) respectively. With the increase in WEMWBS between 8 months and a matched time-point in unvaccinated participants, receipt of a vaccine was statistically associated with a reduction in WEMWBS in a linear model with a coefficient of - 7.6, (95%CI -12.2 ; -2.2, p = 0.006) although there was no significant change in WEMWBS in vaccinated participants themselves (table S3 in the supplement for regression model).

**Figure 3:**
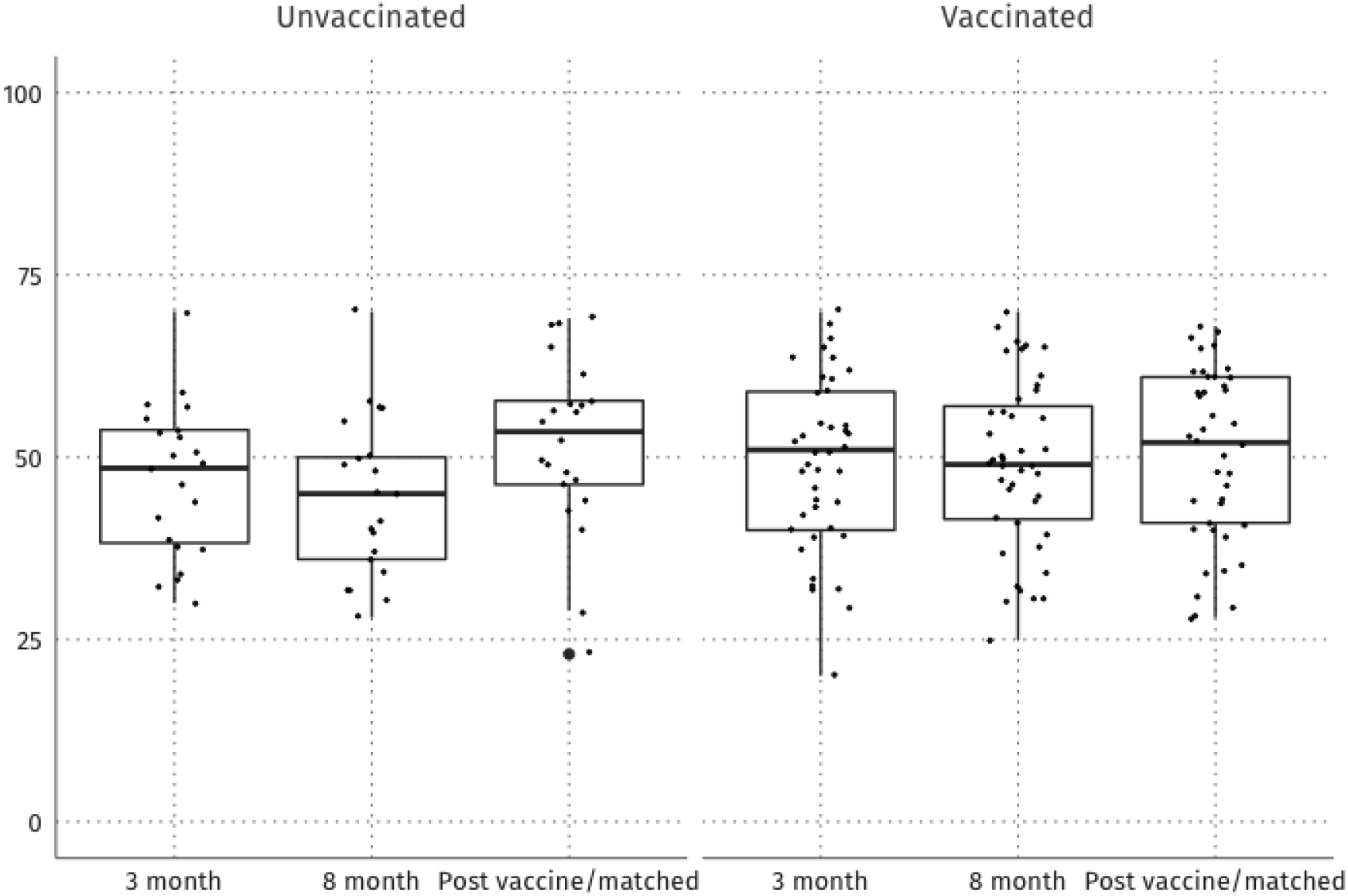
Changes in Edinburgh-Warwick Mental Wellbeing Scale over time.

### Pfizer-BioNTech vs AstraZeneca

Figure S4 compares Pfizer-BioNTech vs AstraZeneca in terms of PCS and MCS. Due to low numbers, this data should be interpreted with caution. However, we found no evidence of any difference in PCS or MCS with either vaccine.

## Discussion

In this cohort of patients with Long Covid following hospitalisation, symptom burden remained high at 8-month follow-up. Importantly, receipt of a vaccine against SARS-CoV-2 was not associated with an increase in symptoms or reduction in quality of life. There was a suggestion of a reduction in symptom burden compared to unvaccinated participants from the same cohort.

Recent studies have demonstrated that in a proportion of individuals have persistent symptoms following COVID-19. Exact estimates vary although have been reported as high as three-quarters in hospitalised patients at 6 months^(2)^ and in up to 10% of community cases^(1)^. The exact cause of what is now commonly referred to as Long Covid remains unclear, although it is likely multi-factorial. Hypotheses include (i) post-viral syndromes given similarities with ME/CFS (ii) persistence of virus or viral proteins within the body e.g. the gut (iii) auto-antibody mediated immunopathology, among others^(5)^. The uncertainty around the cause, especially given the potential immune mediated hypotheses, has led to questions around the potential impact of vaccination to Long Covid symptoms. There is limited evidence base on this topic at present although unpublished literature has suggested both a worsening and improvement of symptoms. Long Covid sufferers are naturally apprehensive to receive an intervention that might worsen symptoms, especially with a perception that the previous infection infers at least some immunity^(3)^. Recent studies of recovered seropositive individuals demonstrate excellent immune response to vaccination compared to seronegative individuals, although they may experience more systemic ‘flu-like’ symptoms (61% in this cohort which all resolved after 48-72 hours), so research that will reduce vaccine hesitancy for Long Covid sufferers is vital.

In this prospective study of patients previously hospitalised with COVID-19 between April to May 2020 the proportion of ongoing symptoms perceived to be related to their admission with coronavirus was over 80%. Numerous multi-system symptoms were reported, although fatigue (61%), breathlessness (50%) and insomnia (38%) were the most common. When comparing vaccinated and unvaccinated participants, there were more symptoms that improved (23.7% vs 15.4%) and less that worsened (5.6% vs 14.3%), a reassuring result, although it is important to note the potential for bias by recall is large in this analysis.

Prior to the vaccination period, participants rated their quality of life as poor compared to population means^(6)^, demonstrating the impact that Long Covid has months after the acute infection. Following vaccination with either the Pfizer-BioNTech or AstraZeneca vaccine there was no significant change in participants self-reported quality of life; which again provides reassurance that vaccination does not reduce quality of life in those hospitalised with prior COVID-19.

For one questionnaire, the WEMWBS, we did identify a statistically significant worsening of the score, but this was driven not by a change in the vaccinated cohort but an increase in score in the unvaccinated cohort. There was no change in the vaccinated cohort over time with respect to this score, and the biological plausibility of a causal relationship between vaccine roll-out and an increase between 8-10 months in this score in the unvaccinated group but a stable response in the vaccinated cohort is questionable.

This study’s strengths include unselected patient recruitment and a robust, longitudinal, assessment of health status, allowing us to confidently measure the impact of any potential vaccine effect on symptom status in a symptomatic cohort. However, the sample size is small, meaning effects on individual symptoms or differences between vaccines cannot be fully explored. Secondly, this is an initially hospitalised cohort so we cannot directly extrapolate to individuals who initial infection did not result in hospitalisation although there is no reason to suggest the effect would be any different. Finally, the most obvious bias is that of blinding and that participants who received vaccination are different in some respects (age, comorbidities) than those who did not. These weaknesses are common to all observational studies, but in particular, symptom recall may well be influenced by receipt of vaccination (a placebo/nocebo effect), and caution must be used in interpreting this data.

In conclusion, receipt of vaccination with either an mRNA or adenoviral vector vaccine to SARS-CoV-2 was not associated in a worsening of symptoms or quality of life in patients with Long Covid. These results should be reassuring for the large numbers of individuals worldwide who have developed persistent symptoms post coronavirus infection and are considering whether to take up the vaccine. When compared to matched unvaccinated participants from the same cohort, those who had received a vaccine actually reported an overall improvement in Long Covid symptoms. Although the sample size is small to make firm conclusions for this observation it is of interest and raises further questions about the aetiology of these symptoms. Individuals with prolonged COVID-19 symptoms should receive vaccinations as suggested by national guidance.

## Data Availability

Data is not available.

## Supplement

**S1:** Regression output for 10 month MCS

| **Characteristic** | **Beta** | **95% CI<sup>†</sup>** | **p-value** |
| --- | --- | --- | --- |
| 8 Month MCS | 0.73 | 0.57, 0.88 | <0.001 |
| Vaccine status |  |  |  |
| Control | — | — |  |
| Vaccinated | 1.6 | -2.7, 5.8 | 0.5 |
| Sex | -1.5 | -5.5, 2.5 | 0.5 |
| Age (18+) | 0.15 | 0.01, 0.30 | 0.043 |
| <sup>†</sup>CI = Confidence Interval |  |  |  |

**S2: Regression output for 10 month PCS**

**S2:** Regression output for 10 month PCS

| **Characteristic** | **Beta** | **95% CI<sup>†</sup>** | **p-value** |
| --- | --- | --- | --- |
| 8 month PCS | 0.93 | 0.80, 1.1 | <0.001 |
| Vaccine status |  |  |  |
| Control | — | — |  |
| Vaccinated | 0.76 | -2.4, 3.9 | 0.6 |
| Sex | -1.0 | -3.9, 2.0 | 0.5 |
| Age (18+) | 0.00 | -0.12, 0.11 | >0.9 |
| <sup>†</sup>CI = Confidence Interval |  |  |  |

**S3:**

**S3:**
| **Characteristic** | **Beta** | **95% CI<sup>†</sup>** | **p-value** |
| --- | --- | --- | --- |
| 8 month WEMWBS | 0.68 | 0.47, 0.89 | <0.001 |
| Vaccine status |  |  |  |
| Control | — | — |  |
| Vaccinated | -7.3 | -12, -2.2 | 0.006 |
| Age (18+) | 0.23 | 0.05, 0.41 | 0.013 |
| Sex | -2.6 | -7.6, 2.4 | 0.3 |
| <sup>†</sup>CI = Confidence Interval |  |  |  |

S4: Pfizer BioNTech vs AstraZeneca vaccines compared across Physical Composite Score

**S4:**
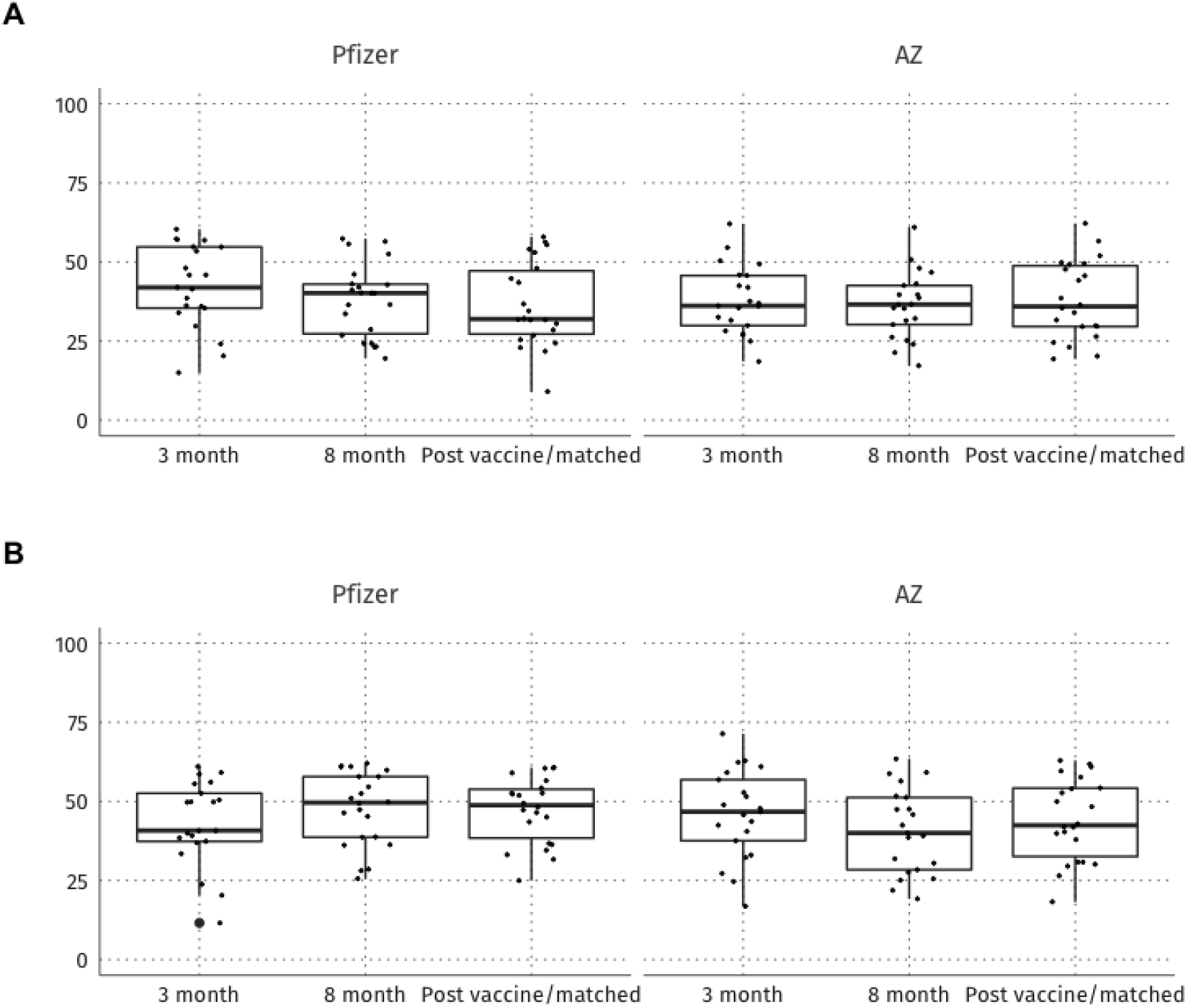
Pfizer BioNTech vs AstraZeneca vaccines compared across Physical Composite Score

